# Clinical validation of an RSV neutralization assay and testing of cross-sectional sera pre- and post- RSV outbreaks from 2021-2023

**DOI:** 10.1101/2024.06.13.24308891

**Authors:** Eli A. Piliper, Jonathan Reed, Alexander L. Greninger

**Affiliations:** Department of Laboratory Medicine and Pathology, University of Washington Medical Center, Seattle, WA, USA; Vaccine and Infectious Disease Division, Fred Hutchinson Cancer Research Center, Seattle, WA, USA

**Author notes:** Corresponding author: Alexander L. Greninger, Department of Laboratory Medicine and Pathology, University of Washington Medical Center 850 Republican St, Seattle, WA 98109, USA.

## Abstract

**Background:** Respiratory syncytial virus (RSV) is a leading cause of acute respiratory infections and hospitalization in infants and the elderly. Newly approved vaccines and the prophylactic antibody nirsevimab have heightened interest in RSV immunologic surveillance, necessitating development of high-throughput assays assessing anti-RSV neutralizing activity.

**Objectives:** We validated an RSV focus-reduction neutralization test (RSV FRNT), a high-throughput, automation-ready neutralization assay using RSV strain A2. The assay was then used to investigate the immunity debt hypothesis for resurgent RSV outbreaks in the 2022-2023 season.

**Study design:** We evaluated precision, sensitivity, specificity, linearity, and accuracy of RSV FRNT using reference sera, contrived specimens, and clinical remnant specimens. RSV neutralizing activity of remnant serum specimens, sampled for HSV-1/2 antibody testing during and after the COVID-19 pandemic (February and September 2022 & 2023), was measured and correlated with concurrent trends in RSV prevalence.

**Results:** RSV FRNT was shown to be accurate, generating reference serum neutralizing titers within 2-fold of established assays, with a linear analytical measurement range between 20 to 4,860 ND50 and ND80 units. Neutralizing activity measured with the assay was positively correlated with antibody titer determined via indirect ELISA (ρ = 1.0, p = 0.0014). Among individuals sampled within 3 months of RSV PCR test, RSV positives had a 9.14-fold higher geometric mean neutralizing titer (GMT) relative to RSV PCR negatives (p = 0.09). There was no difference in geometric mean anti-RSV neutralizing titers between sera sampled pre- and post-2023 RSV outbreak (p = 0.93).

**Conclusions:** We validated a high-throughput assay for assessing anti-RSV neutralizing activity and found no significant difference in neutralizing titers between pre- and post-pandemic outbreak specimens.

**Highlights:**

- We report the full clinical validation of an RSV neutralization assay
- No evidence of immune debt found in Washington State RSV 2022-2023 outbreak
- Our assay offers high throughput testing applicable to vaccine/drug studies

## Introduction

RSV is one of the most common causes of lower respiratory tract infections worldwide, infecting approximately 64 million people and causing at least 160,000 deaths and 3 million hospitalizations each year [1,2]. 2023 has seen multiple revolutions in RSV disease prevention, including the FDA approval of two vaccines for older adults and pregnant individuals as well as the prophylactic monoclonal antibody (mAb) nirsevimab for prevention of RSV lower respiratory tract disease in neonates and infants [3,4]. In addition, the past two years have seen global resurgence of aseasonal RSV infections thought to be related to limited circulation and immunity debt attributed to the COVID-19 pandemic [3,5–8]. These developments necessitate monitoring population immunity and vaccine immunogenicity, both best assessed by measuring neutralizing antibodies against RSV [9–12]. RSV neutralizing antibody titer is a major correlate of protection, with higher neutralizing titers being associated with lower viral load and reduced risk of severe RSV-induced disease [13–18].

The standard for neutralization assays, the plaque-reduction neutralization test (PRNT), requires 3-5 days of incubation time and is less amenable to high-throughput applications [12]. An alternative is the focus-reduction neutralization test (FRNT), which often requires less sample, reagent volume, and incubation time (26-30 hours) to measure neutralizing titers. A RSV FRNT assay can be performed in a 96-well plate format portable to liquid handlers for higher throughput and less inter- and intra-assay variability [19–22]. Strain-specific reporter viruses are utilized by some FRNTs to visualize foci but require development of a recombinant virus, potentially delaying efforts to implement contemporary strains in testing. A FRNT using chromogen-conjugated secondary antibodies to visualize foci is easy to implement, allowing scalability for high-throughput applications such as vaccine evaluation, population immunity monitoring, and testing for vaccine- and nirsevimab-resistant strains [23].

Here, we describe the validation of a RSV FRNT, an automation-ready, high-throughput, standardized method for assessing serum neutralizing activity against RSV in clinical and research settings. In addition, we use the RSV FRNT to monitor population immunity against RSV in remnant serum specimens taken before, during, and after the 2022-2023 RSV outbreak.

## Materials and Methods

### Virus and cell line

Assays were run with RSV A2 as the challenge virus (ATCC, VR-1516), unless otherwise specified. RSV B strain WV/14617/85 (ATCC, VR-1400) challenge experiments are described in Supplemental Methods. VeroE6 cells were seeded in DMEM-10 in transparent 96-well plates (Corning, 3585) at 10,000 cells/well and grown for 24 hours prior to infection.

### Clinical specimens

Plasma or sera from patients with a history of positive RT-PCR test for RSV or influenza virus since October 1, 2023 were obtained from University of Washington (UW) Medicine patients that were sampled during peak RSV incidence (mid-November 2023 to January 2024) and no later than 40 days following PCR testing. Remnant serology specimens sent for rubella IgG testing were obtained from UW Virology. Cross-sectional sera were obtained from remnant specimens sent for HSV-1/2 Western Blot testing in February 2022, July 2022, February 2023, and July 2023 (n=47 for each timepoint). This study was approved by the UW Medicine Institutional Review Board with consent waiver (STUDY00010205). Individual line-item data and metadata is provided in Table S1. For individuals with recorded RSV PCR tests, time delta (in days) between serum collection and RSV PCR test date is shown in Figure S1.

### Measurement of RSV neutralizing activity

#### Microneutralization

All sera were heat inactivated (HI) at 56 ° C for 30 minutes prior to testing. HI serum was diluted initially 10-fold by combining 10.5 µL HI serum with 94.5 µL DMEM-10. This was followed by five 3-fold dilutions by serially transferring 35 µL of diluted serum into 70 µL of DMEM-10, resulting in a final dilution series from 10-fold to 2430-fold and a final volume of 70 µL for each dilution. These serial dilutions of serum were diluted an additional two-fold to a final volume of 140 µL with 70 µL of RSV A2, diluted to form a final of 120-250 ffu/well in the assay, resulting in a final dilution series of 20-fold to 4,860-fold. Serum-virus mixtures were incubated at 37° C 5% CO_2_ for 1 h. 96-well plates seeded with 10,000 VeroE6 cells/well were infected with 50 µL/well of serum-virus mixture in duplicate. Plates were incubated at 37° C 5% CO_2_ for 26 – 30 hours before fixation. Exogenous complement, typically[13,24] used to boost neutralizing signal from low-titer sera, was not used in our RSV FRNT; instead, samples with neutralizing titers below the lower limit of quantitation were retested at a lower, 5-fold initial dilution to obtain reportable ND50 and ND80 values within assay analytical measurement range (20-4,860).

#### Focus-forming assay

Assay plates were fixed for 60 minutes at room temperature (approximately 21-25 ° C) with 25 µL (1/2 well volume) of 4% formaldehyde prepared in DPBS with 100 mg/L calcium and 100 mg/L magnesium.

After fixation, plates were washed three times with 150 µL room-temperature 1xPBS and incubated for 2 h at room temperature with 50 µL/well of mouse mAb 131-2G against RSV-A2 F-protein (Sigma-Aldrich, MAB8582) diluted 4000-fold in FFA staining buffer (composed of 1x PBS, 1 mg/mL saponin, and 0.1% IgG-free BSA). Plates were then washed three times with 150 µL room-temperature FFA wash buffer (composed of 1x PBS and 0.05% Triton X-100). Plates were incubated for 1 h at room temperature with 50 µL/well of peroxidase-conjugated secondary antibody (Bethyl Laboratories, A90216P) diluted 4000-fold in FFA staining buffer. Following secondary incubation plates were then washed three times with 150 µL room-temperature FFA wash buffer. Plates were developed with 50 µL TrueBlue peroxidase substrate (KPL, 5510-0054) for 15 minutes at room temperature. Development was stopped by washing three times with 150 µL room-temperature deionized water, and the plates were dried for 10-15 minutes to minimize visual artifacts from liquid.

#### Imaging and counting

Plates were scanned and counted on the S6 Universal M2 ImmunoSpot analyzer (Cellular Technology Ltd, Cleveland, OH). See supplementary information for counting settings. Counts were used to determine percent foci reduction at each dilution. For each specimen, the response between percent foci reduction and serum dilution was modeled via 4-parameter logistic regression (Supplementary Material), which was used to obtain ND50 and ND80 values.

## Results

### Assay Accuracy

To determine if RSV FRNT results closely reflected binding antibody titers, we tested remnant rubella serology specimens and contrived serum samples via RSV FRNT and anti-RSV IgG indirect ELISA testing (Supplementary Methods), with the expectation that specimens with higher ND50 values would have higher anti-RSV antibody titers by ELISA [25]. We prepared a “linearity panel” of six contrived specimens derived from NR-21973 reference serum with theoretical ND50 values ranging from 15-14,982 (Supplemental Methods). We tested these alongside 22 remnant rubella serology specimens. RSV FRNT ND50 values of linearity panel members were positively correlated (ρ = 1, p = 0.0014) with the level of RSV-reactive antibodies detected by ELISA (Figure 1A). All 22 rubella serology specimens tested positive for neutralizing antibodies by FRNT, with ND50 values spanning the full range of measurable ND50 values between the lowest to highest serum dilutions (20-4,860). Similarly, 21 of 22 rubella serology specimens were positive for RSV-reactive antibodies by ELISA based on the manufacturer’s cutoff. The one remnant rubella serology specimen that fell just below the ELISA positive cutoff had an ND50 of 864. An Ig-depleted control exhibited an ND50 < 20 in the RSV FRNT assay and fell below the ELISA positive cutoff (Figure 1A).

**Figure 1.**
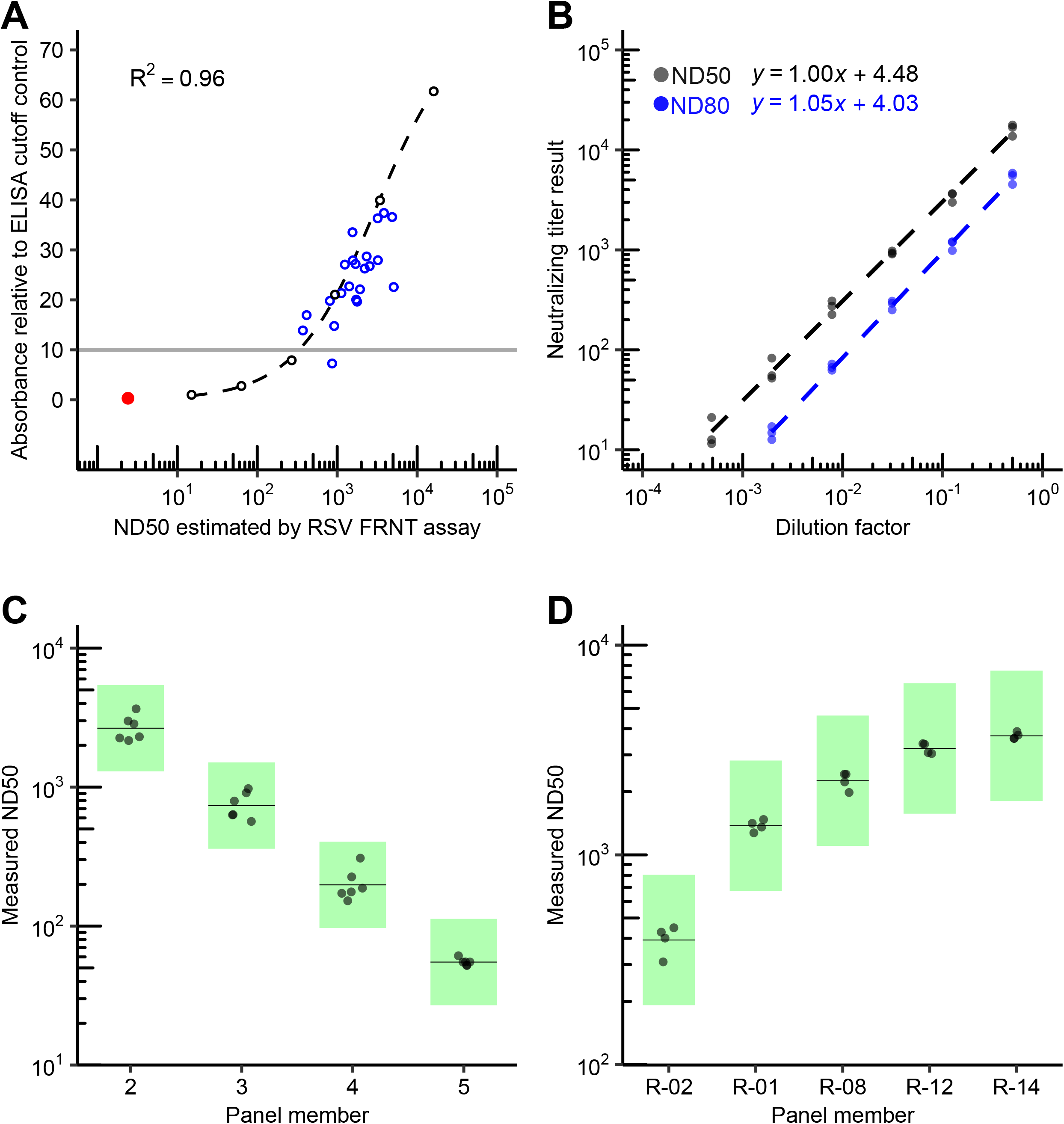
The RSV FRNT assay results are strongly correlated with ELISA measurements and meet our linearity and precision requirements. A) Shown are the results of RSV-binding antibody measurement by ELISA of the linearity panel specimens (black open circles; see Assay Accuracy), rubella serology remnant specimens (blue open circles), and the Ig-depleted negative control (red closed circle, NR-49447). Dashed black line is the 4-parameter logistic curve fit of the ELISA results from the linearity panel only (R^2^ = 0.96). Sera was measured in duplicate, normalized relative to the assay cutoff control (relative absorbance of ten), and plotted as average values. The standard deviation of the measurements is plotted as a black line range. In most cases, the standard deviation was smaller than the diameter of the plotted points obscuring the standard deviation line range. Values with relative absorbance less than nine are considered negative since they fall below the cutoff. Values with relative absorbance between 9 and 11 are inconclusive. Values with relative absorbance greater than 11 are positive for RSV-reactive antibodies. B) RSV FRNT ND50 and ND80 values from testing of the linearity panel. The range of mean ND50 results include from 15.1 to 16,093.2 and the range of mean ND80 results include from 14.9 to 5308.7. The dashed lines (black for ND50 and blue for ND80) are the best fit from the linear regression analysis. C) Linearity panel specimens (the same used in panel A and B) ranging in expected ND50 values from 55 to 14,982, were measured in duplicate over 3 days. Every sample exhibited GCV < 37%, meeting our acceptance criteria for precision. Green boxes demarcate boundaries of expected variation for a GCV of 37%. Solid lines represent mean ND50 for panel member. D) Five serum specimens from rubella remnant testing with known ND50s/ND80s spanning the AMR of the assay were selected and tested in duplicate over two days. Every sample exhibited GCV < 37%, meeting our acceptance criteria for precision. Green boxes demarcate boundaries of expected variation for a GCV of 37%. Solid lines represent mean ND50 for panel member.

We next compared ND50 values of our RSV FRNT assay with an independent reference assay on commercially available sera and standardized results o the First International Standard for Antiserum to Respiratory Syncytial Virus (NIBSC code: 16/284) [26]. RSV FRNT results were within 2-fold of previously published values (Table 1) [26]. FRNT results were also within 2-fold of proposed reference measurements when using RSV strain B WV/14617/85 (Table 2) 6/13/2024 10:32:00 AM. The conversion factor for RSV A2 and RSV B FRNT ND50 units to IU/mL was 0.37 and 0.91, respectively.

**Table 1:** ND50 values, expressed in IU/mL, obtained by the RSV FRNT assay using RSV A2 as the challenge virus are within 2-fold of independent published estimates.

| sample | n | ND50 RSV FRNT (IU/mL)<sup>a,c</sup> | ND50 reference (IU/mL)<sup>b,c</sup> | ratio of ND50 RSV FRNT / reference |
| --- | --- | --- | --- | --- |
| NR-21973 | 5 | 9,539 | 6,830 | 1.4 |
| NR-4020 | 5 | 1,392 | 1,236 | 1.1 |
| NR-4021 | 6 | 4,154 | 4,404 | 0.9 |
| NR-4022 | 6 | 898 | 706 | 1.3 |
| NR-4023 | 12 | 1,114 | 625 | 1.8 |

<sup>a</sup>NIBSC 16/284 standard was tested twelve times with RSV A2, with geometric mean 5404.8. To convert ND50 values to IU/mL, ND50 values obtained in the RSV FRNT assay are multiplied by 0.37.
<sup>b</sup>Reference values were taken from an independent publication, which used an IU/mL conversion factor of 0.88[26].
<sup>c</sup>Data represents the geometric mean.

**Table 2:** ND50 values, expressed in IU/mL, obtained by the RSV FRNT assay using RSV B WV/14617/85 as challenge virus are within 2-fold of independent published estimates.

| sample | n | ND50 RSV FRNT (IU/mL)<sup>a,c</sup> | ND50 reference (IU/mL)<sup>b,c</sup> | ratio of ND50 RSV FRNT / reference |
| --- | --- | --- | --- | --- |
| NR-21973 | 6 | 1,107 | 1,005 | 1.1 |
| NR-4022 | 6 | 13,738 | 12,059 | 1.1 |

<sup>a</sup>NIBSC 16/284 standard was tested 6 times, with geometric mean 2,195.8. To convert ND50 values to IU/mL, ND50 values obtained in the RSV FRNT assay are multiplied by 0.91. In the reference article, mean conversion factor across all assays reported was 1.1; the reference did not report conversion factor per-assay[27].
<sup>b</sup>Reference values obtained from independent publication (no conversion factor provided)[27].
<sup>c</sup>Data represents the geometric mean.

### Assay Linearity, Precision, Reportable Range, and Limits of Quantitation

We next analyzed ND50 and ND80 results from the linearity panel by linear regression. Linearity panel ND50 and ND80 results within or near the assay lowest and highest dilutions (20-4,860) were strongly linear (R^2^ > 0.9), with slopes of 1.00 and 1.05, respectively. This met our assay acceptance criteria of R^2^ > 0.9 and slope between 0.9 and 1.1 (Figure 1B, Table S2).

We established our maximum acceptable geometric coefficient of variation (GCV) to be 37%, based on CLSI and FDA requirements [28], and precision criteria of similar assays [29–39] (see Supplemental Material for further discussion). To estimate intra-assay, inter-assay, and overall imprecision, we ran four linearity panel specimens (in duplicate over 3 days, Figure 1C) and five rubella serology specimens (in duplicate over 2 days, Figure 1D). No specimen, clinical or contrived, exhibited intra-assay, inter-assay, or overall GCV above 37% (Table S3 and S4). Measurements on reference serum using RSV B WV/14617/85 as a challenge virus also had GCV < 37% (Table S5).

Per the precision analysis, ND50 and ND80 GCV was < 37% near the minimum and maximum dilutions (20 and 4,860), and assay measurements were linear through this range. Thus, the lower limit of quantitation and upper limit of quantitation of the assay were determined to be 20 and 4,860, respectively, for both ND50 and ND80. To verify values above the analytical measurement range, we prepared four dilutions of NR-21973 (ND50 29,963) and recovered values within 1.2-fold of original ND50 values (Supplementary Material, Table S6).

### Comparison of 2021-2023 RSV neutralization titers associated with RSV outbreaks

To investigate the role of population-level immune debt in the winter 2022 RSV outbreak, we determined the geometric mean titer (GMT, geometric mean of ND50 measurements) of a repeated cross-sectional collection of 193 remnant sera sent for clinical HSV western blot testing sampled across four discrete timepoints between 2022-2023: after 2021 RSV outbreak (February 2022), before (August 2022) and after (February 2022) 2022 RSV-outbreak, and before 2023 RSV outbreak (September 2023, Figure 2). We found no significant difference in GMT or median geometric titer between any combination of the four timepoints (one-way ANOVA, p = 0.86). Moreover, samples collected before and after the 2022 RSV outbreak exhibited no significant difference in GMT (Wilcoxon ranked-sum test, p = 0.68, Figure 2). In addition to the cross-sectional remnant HSV sera, we tested RSV neutralizing titers of a separate group of remnant HSV serology serum specimens from 26 individuals who tested PCR positive for RSV after serum collection (by 25 – 463 days), referred to as future-RSV-PCR-positives. The overall GMT from these individuals was not significantly different from the cross-sectional sera for which RSV PCR testing data was not available (two-sided t-test, p = 0.24, Figure 3).

**Figure 2.**
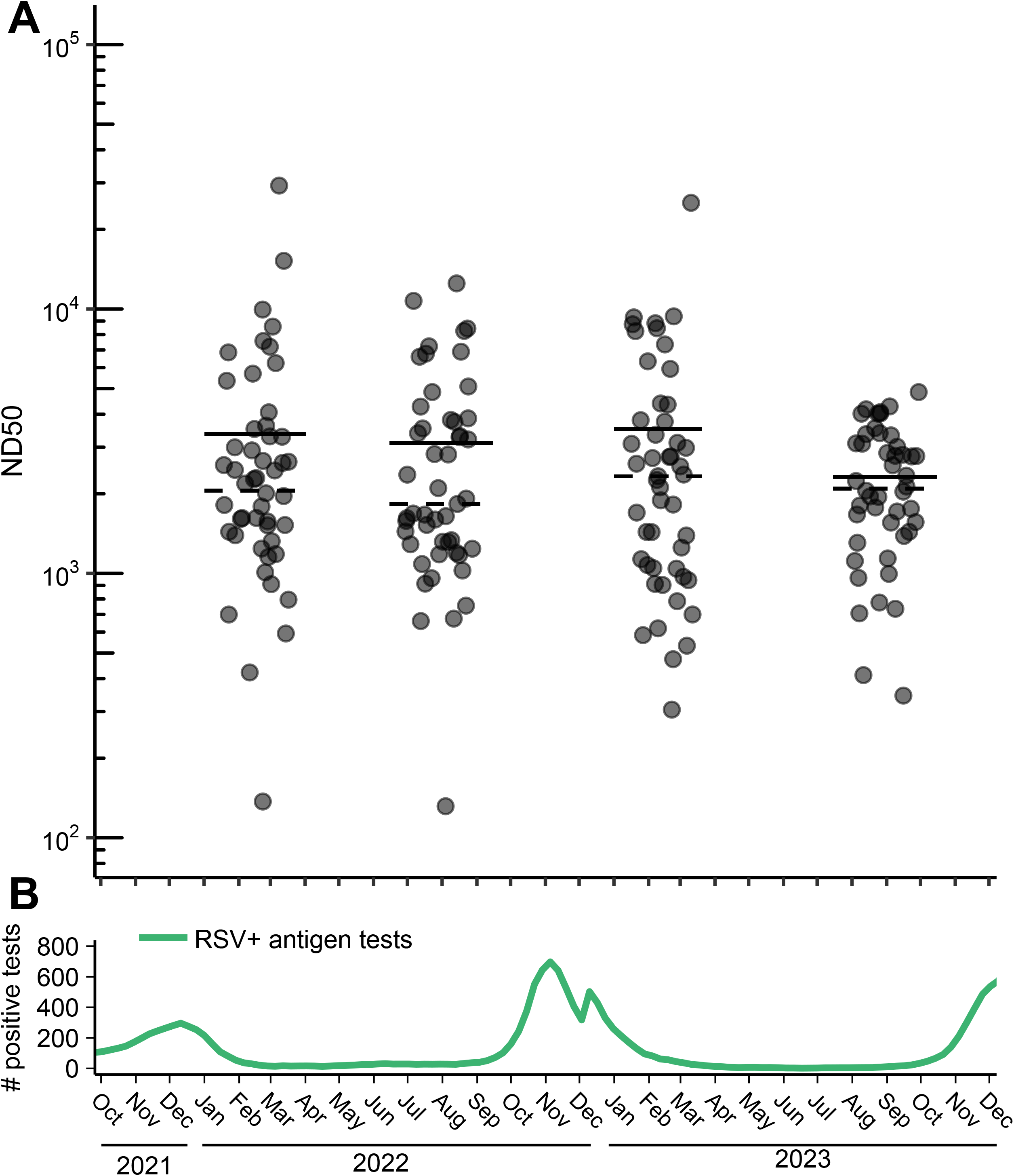
Distribution of neutralizing titers among random population samples collected from late 2021 to early 2024. A) The ND50 neutralizing titer results of HSV serology remnant serum specimens from individuals sampled around February 2022, August 2022, February 2023, and September 2023, with date of serum collection on x-axis. For each cross-sectional group, dashed and solid lines represent geometric median and geometric mean, respectively. B) Number of positive RSV antigen tests across time. Data obtained from CDC Washington State trends[62], reported as 5 week average. Log_2_-transformed data was analyzed, using a two-sided student’s T-test to compare groups pairwise.

**Figure 3.**
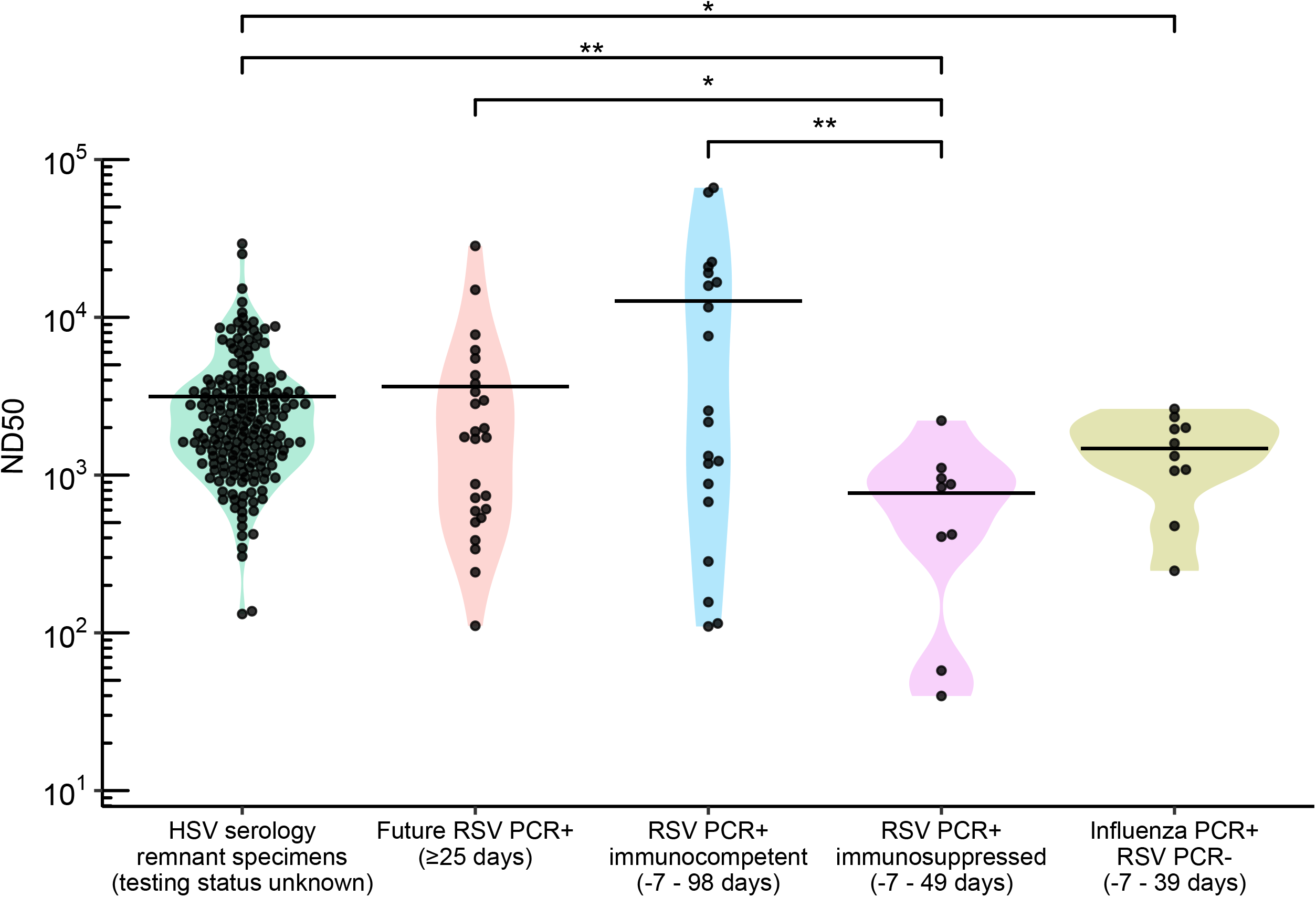
Distribution of neutralizing titers among RSV negative, positive, and future-infected individuals relative to a random population sample. ND50 neutralizing titer for individuals sampled from October 2021 to February 2024. Solid lines represent geometric mean. HSV serology remnant specimens: a random population sample of remnant sera sent for clinical HSV remnant testing, collected from February 2021 to September 2023, with unknown testing status for RSV. Future RSV-infected: HSV remnant samples (not part of the HSV serology remnant specimen group above) from individuals who tested positive for RSV 25 or more days after serum collection date. RSV PCR+, immunocompetent: emergency department (ED) patients PCR-positive for RSV 98 days before to 7 days after serum collection. RSV PCR+, immunosuppressed: ED patients PCR-positive for RSV 49 days before to 7 days after serum collection, with medical status of immunosuppression at time of test/collection. Influenza+, RSV-: ED patients with no recorded RSV-positive or immunosuppression status who tested PCR-positive for influenza 39 days before to 7 days after serum collection. Log_2_-transformed data was analyzed, using a two-sided student’s T-test to compare groups pairwise.

To further profile neutralizing titers in individuals testing PCR positive or negative for RSV, we collected remnant sera from UW Medicine patients who had recently (within 3 months of sampling) tested PCR positive for RSV (n=29) or PCR negative for RSV and positive for influenza virus (n=10) during the 2023-2024 respiratory season (Figure 3). The PCR positive RSV group included 9 immunosuppressed individuals based on chart review and were separated into their own group for this analysis. Population GMT (combined cross-sectional HSV serology remnant specimens sets described above) was 4.2-fold lower than immunocompetent RSV PCR positive individuals (two-sided t-test, p = 0.51), 4.1-fold higher than that of RSV PCR positive immunosuppressed individuals (p = 0.008), and 2.14-fold higher than that of influenza PCR-positive, RSV PCR-negative individuals (p = 0.04). The future-RSV-PCR-positive samples had a GMT 4.7-fold higher than PCR positive, immunosuppressed individuals (p = 0.03) and 3.5-fold lower than PCR positive individuals without immunosuppression (p = 0.24). Immunocompetent RSV PCR positive individuals had a GMT 17.4-fold higher than that of RSV PCR positive immunosuppressed individuals (p = 0.007), and 9.14-fold higher than Influenza PCR-positive, RSV PCR-negative individuals (p= 0.09). Thus, the measurements obtained by RSV FRNT align with patient medical background and can represent individuals ranging from the recently infected and immunocompetent to the immunosuppressed.

## Discussion

Here, we designed and validated a high-throughput RSV neutralization assay amenable to clinical testing, clinical trials, and seroprevalence studies. As a precise, sensitive, and linear assay standardizable to the NIBSC 16/284 RSV neutralizing reference material, RSV FRNT is an alternative to reporter virus methods, allowing for similar throughput, quantification, and precision unlimited by development of recombinant viruses. The potential to use wild-type strains as challenge viruses also enables monitoring for nirsevimab resistance and vaccine evasion by RSV, which are potential concerns given the antigenic variability of the RSV F-protein [40–42], continued vaccine roll-out[4], and recent identification of RSV strains resistant to nirsevimab[43,44].

Our assay had similar if not improved precision compared to other assays [29–34]. Our RSV FRNT differs from most other assays with a somewhat lower upper AMR limit of only 4,860 (ND50 or ND80) [45]. Given that only 2.6% (n=6/226) of tested clinical specimens exhibit a neutralizing titer (NT)> 10,000, we found the increased throughput more desirable. We also standardized our RSV FRNT to the First International Standard for Antiserum to Respiratory Syncytial Virus strains A and B, which has been reported for select assays [24]. Our RSV A2 conversion factor of 0.37 is very similar to the 0.38 conversion factor used in a 2021 RSV vaccine clinical trial [46]. In contrast, the RSV A conversion factor from another study was 0.88 [26]. The higher conversion factor is most likely explained by key differences in assay methodology, such as challenge virus, target cell line, and assay readout [24,45]. Our RSV B conversion factor was 0.91, similar to the average conversion factor of 1.1 calculated from NIBSC [27].

We found no significant difference in neutralizing titer between individuals serums sampled pre- and post-RSV 2022 outbreak, unlike earlier studies which found evidence of reduced anti-RSV neutralizing titer during the COVID-19 pandemic [11,47]. In agreement with our findings, a 2022 population-immunosurveillance study using methods similar to ours also could not detect waning antibody immune response prior to RSV resurgence [48].

Moreover, a recent study attributed approximately two-thirds of the increase in RSV cases to significant increases in volume of RSV testing [49]. Other possible theories behind post-pandemic RSV resurgence, such as SARS-CoV-2-induced immune dysregulation [50–53], changes in population-wide health-seeking behavior [54], and loosening of public health safety measures [8] should also be investigated to inform future vaccination and immunotherapeutic development.

Limitations of this study include its cross-sectional design and convenience sampling of remnant clinical specimens for validation. Measurement of pre-and post-outbreak RSV neutralizing titers were limited by the sample size (n=227). Our sample group lacked infants (n=0) and had a limited number of elderly individuals (n=22, 12.4%), both of which are especially vulnerable to lowered anti-RSV immunity and severe RSV symptoms[2,57,58]. Patients with no reported RSV infection might have experienced unreported RSV infection from the smaller outbreak of winter 2021, potentially inflating neutralizing titers and confounding comparison of pre- and post-2022 outbreak populations. RSV strains A2 and B WV/14617/85, classically used for FRNT and PRNT validation[24], were used instead of more contemporary and/or clinical isolates in circulation during the 2022-2023 pandemic[59–61]. Differences in sample handling may have influenced measurements. For example, we noticed that GMT across sera of RSV PCR-positive individuals (without immunosuppression) sourced from one hospital trended lower compared to those from the two other hospitals used in the study by 4.43-fold and 2.55-fold respectively, although this difference did not reach statistical significance.

Our findings establish the RSV FRNT assay as a high-throughput, accurate neutralization assay sufficiently sensitive to distinguish immunosuppressed and seroconverted individuals from the general population, a diagnostic necessity given the upcoming release of the RSV vaccine. Our immunosurveillance study using this assay found no evidence of immune debt, underlining the uncertainty behind drivers of RSV epidemiology and RSV antibody dynamics, an ever-more concerning gap in knowledge considering the eventuality of the next viral pandemic.

## Data Availability

All data produced in the present study are available upon reasonable request to the authors

## Acknowledgments

We thank Dr. Stephanie Goya for insightful discussion and writing assistance. This research received no specific funding and was supported by departmental funds.

