## Supplementary material for "Clinical validation of an RSV neutralization assay and testing of cross-sectional sera pre- and post- RSV outbreaks from 2021-2023": Figure S1

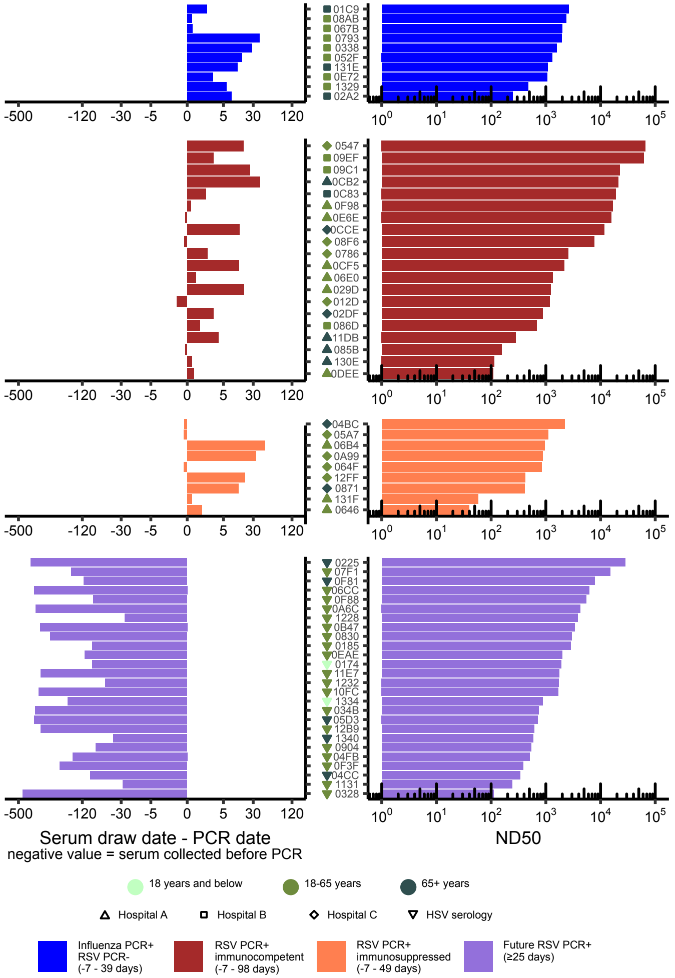


**Figure S1: Time delta between serum and RSV PCR test, with ND50 values.** Hospital names were anonymized and patients given random hexadecimal identifiers. Left: Difference in days between serum collection and RSV PCR date, calculated as (serum collection date - PCR test date). Negative values indicate PCR tests taken after serum draw. Right: ND50 measurements. Influenza PCR+, RSV PCR-: emergency department (ED) patients with no recorded RSV-PCR-positive or immunosuppression status, who tested PCR-positive for influenza from 39 days before to 7 days after serum collection. RSV+, immunocompetent: ED patients PCR-positive for RSV from 98 days before to 7 days after serum collection date. RSV+, immunosuppressed: ED patients PCR-positive for RSV from 49 days before to 7 days after serum collection date, with medical status of immunosuppression at time of serum draw. Future RSV PCR+: individuals with serum drawn 25 days or more before RSV-positive PCR test.

### Figure S1: Time delta between serum and RSV
