## Supplemental Material for "Clinical validation of an RSV neutralization assay and testing of cross-sectional sera pre- and post- RSV outbreaks from 2021-2023"

*Supplemental Results*

*Precision, Reportable Range and Limits of Quantitation*

General acceptance criteria for assay imprecision range, formulated from CLSI and FDA standards, require a maximum arithmetic coefficient of variation (CV) of 30% around the mean measurement value [1]. Given the log-normal distribution of neutralization data and the more variable nature of cell-based assays, other similar methodologies utilize geometric coefficient of variation (GCV), with acceptable GCV maxima ranging from 20% to 50% [2–13]. A geometric standard deviation of 1.43 (GCV of 37%) was selected as an imprecision threshold for the RSV FRNT assay to ensure that the true population mean is at least within 2-fold of the sample mean 95% of the time.

To estimate intra-assay, inter-assay, and overall within-lab imprecision of RSV FRNT, we measured 4 samples belonging to the linearity panel and tested them in duplicate over 3 days (Table S3). Rubella serology remnant specimens were also tested in duplicate over 2 days (Table S4). Imprecision of ND50 and ND80 values were estimated via ANOVA analysis. Contrived specimens had ND50 values with average total GCV of 20.7%, and ND80 values with average total GCV of 22.2% (Figure 1C, Table S3). Clinical specimens had ND50 values with average total GCV of 8.1% and ND80 values with average total GCV of 8.5% (Figure 1D, Table S4). No specimen, contrived or clinical, exhibited a mean GCV > 37% for overall imprecision, meeting our precision criterion of GCV < 37% (Tables S3 and S4). Neutralizing titers obtained from tests using RSV B WV/14617/85 also had overall GCV < 37% (Table S5).

To gauge if ND50 measurements within the clinical reportable range (CRR), achieved via specimen pre-dilution, were representative of original titers, we prepared four pre-dilutions of NR-21973 (ND50 29,963) to have expected ND50s of 3,745, 936, 234, and 59, respectively and ran them in triplicate. When accounting for pre-dilution, all samples had CRR ND50s within 1.2-fold of the original value of 29,963 (Table S6).

*Analytical sensitivity*

The limit of detection (LoD) of the assay is the minimum percent inhibition required that can be distinguished from an uninhibited control. First, the limit of blank was determined to be 14.5 % inhibition (Table S8) based on the mean percent inhibition and standard deviation of negative control wells receiving virus with no serum, which were 0.00% and 8.80% respectively (Table S7). The LoD was determined to be 28.2% inhibition (Table S8) based on the variation of NR-4023 diluted down to near-LoB level, which was 8.30% (Table S7). Since 50% and 80% inhibition are well above the LoD of the inhibition assay, ND50 and ND80 measurements within AMR are distinguishable from blank.

*RSV strain B Neutralizing titers*

To evaluate the compatibility of RSV FRNT with a B-strain challenge virus, we measured reference sera NR-4022 and NR-21973, along with the NIBSC-16/284 international standard, in 6 replicates across 3 runs using RSV strain B WV/14617/85. All other assay methods remained unchanged. NR-21973 and NR-4022 mean ND50 measurements were within 2-fold of references (1,107 vs 1,005, and 13,738 vs 12,059 respectively[14]; Table 2). Intraassay, interassay, and overall GCV was below 37%, meeting our precision acceptance criteria (Table S5).

*Analytical specificity*

Because RSV seroprevalence is high [15,16] and reference serum unreactive to RSV has not been reported, we used commercially available Ig-depleted pooled serum specimens (NR-49947 and S5393[17,18]) to evaluate analytical specificity and confirm that antibodies were necessary for statistically significant neutralizing activity. Both sera exhibited ND50 measurements below the lower AMR bound of 20 (2.9 and 3.9 respectively, data not shown) and the percent inhibition at the lowest dilution tested fell below the assay limit of detection (<28.2%, see Analytical sensitivity and accuracy). Together, these data demonstrate that this assay can specifically detect antibody-induced neutralization of RSV.

***Supplemental Methods***

*Growth of challenge virus*

RSV strain A2 was grown in Hep-2 cells (ATCC CCL-23), while RSV strain B WV/14617/85 was grown in VeroE6 cells (ATCC, CCL-1587). The following procedure was used for growing both strains: Cells were seeded in T-75 flasks (Corning, 430614U) to reach ~80-90% confluency overnight. On the day of infection, cells were first washed with DPBS (Gibco, 14190-144). Virus was then resuspended in DMEM-2 (for VeroE6 cells) or MEM-2 (for Hep2 cells). DMEM-2 was made from high-glucose DMEM with glutamax and sodium pyruvate (Thermofisher, 10569010), 10mM HEPES (Thermofisher, 15630080), 1% penicillin-streptomycin (Thermofisher, 15140122), and 2% heat-inactivated FBS (Thermofisher, A3840001). MEM-2 was made from MEM with L-glutamine (Thermofisher 11095080), 10mM HEPES (Thermofisher, 15630080), 1% penicillin-streptomycin (Thermofisher, 15140122), and 2% heat-inactivated FBS (Thermofisher, A3840001). Resuspended virus was then added to cells at 0.01 to 0.1 MOI. Flasks were incubated at 37°C 5% CO_2_ until 80% or more of the monolayer was either destroyed or manifesting syncytia (3-7 days). Virus was harvested by scraping off cells into flask supernatant. This material was centrifuged at 4°C for 5 minutes at 200xg. Following removal of supernatant from the cell pellet, the pellet was resuspended in media and subjected to three freeze-thaws to release cell-bound virions. Media from the freeze-thawed pellet was combined with original supernatant and mixed thoroughly. This mixture was diluted 2-fold with sterile-filtered 50% w/v sucrose (JT Baker, 4097-04, resuspended in DPBS) to produce virus stock with 25% w/v sucrose to reduce degradation of virion infectivity in storage[19]. This mixture was aliquoted into 200 µL volumes and stored in -80C. Aliquots were only freeze-thawed once before being discarded to minimize titer variation from viral degradation.

VeroE6 cells (ATCC, CCL-1587) for neutralization assays were maintained below 30 passages and grown in DMEM-10, consisting of high-glucose DMEM with glutamax and sodium pyruvate (Thermofisher, 10569010), 10mM HEPES (Thermofisher, 15630080), 1% penicillin-streptomycin (Thermofisher, 15140122), and 10% heat-inactivated FBS (Thermofisher, A3840001).

*Immunospot counter settings*

Plates were scanned and counted on the S6 Universal M2 ImmunoSpot analyzer (Cellular Technology Ltd, Cleveland, OH). Initial sensitivity/detection settings were calibrated via the Smart Well™ feature trained on no-serum, virus-only control wells. Counted well area was reduced to 80% without normalization to reduce spurious counting of well shadows. Spot separation was set to 25 to avoid counting larger, single foci as multiple foci. Background balance was set to 0 to reduce counting background artifacts in the form of diffuse, low contrast putative foci. Sample sensitivity was held between 197 and 210 and adjusted to ensure that virus only control well counts numbered 150-250 and that foci counted in no-serum, no virus wells, typically from fibers or plate markings, were ≤ 5 foci.

*Construction of contrived specimens***.**

Ig-depleted pooled serum specimens (NR-49447, BEI Resources, and S5393, Sigma-Aldrich) were used as negative controls for assay validation, as both exhibited neutralizing activity below the LoD. Anti-RSV reference serum NR-21973 was obtained from BEI Resources, determined to have an ND50 of 29,963 by the RSV FRNT, was diluted 2-fold, then serially diluted 4-fold with DMEM-10 to produce six contrived specimens along a spectrum of theoretical ND50s (14982, 3745, 936, 234, 59, 15). These contrived specimens were split into single-use aliquots to minimize freeze-thaw effect and stored at -80°C.

*Indirect ELISA*

Samples were measured with anti-RSV Human IgG ELISA (Abcom ab108765) per manufacturer protocol, using included 96-well plates. Plate absorbance was measured at 450 nm and 620 nm immediately after adding stop solution, and optical density was background-corrected by calculating the difference of 450 nm and 620 nm. Substrate blanks, negative controls, cut-off controls, and positive controls were checked to be within manufacturer criteria. Per manufacturer instructions, mean cutoff was calculated from both cut-off control wells. Normalized absorbance relative to the cutoff was calculated by dividing sample absorbance by mean cutoff control absorbance, then multiplying by 10. Samples with normalized absorbance of 11 or above were considered positive.

*Data analysis*

Curve fitting for RSV FRNT data was performed using the lmfit python package using the Nelder-Mead method. Constraints used were a hill slope between -1.2 and -0.7 (initial guess = -0.8), upper limit between 0.95 and 1.05 (initial guess = 1), lower limit between -0.05 and 0.05 (initial guess = 0), and inflection point between 0 and infinity (initial guess = 50). Code used for analysis can be found here: <https://github.com/greninger-lab/rsv_neut_utils>.

Curve fitting for comparing RSV FRNT ND50 with ELISA absorbance was performed with R package dr4pl[20,21], using the Nelder-Mead method. Constraints used were a hill slope between 0.3 and 1.2 (initial guess = 0.5), upper limit between 0 and 1000 (initial guess = 200), lower limit between -100 and 100 (initial guess = -1), and inflection point between negative infinity and 15,000 (initial guess = 10,000).
